# Objective and Subjective COVID-19 Vaccine Reactogenicity by Age and Vaccine Manufacturer

**DOI:** 10.1101/2021.04.29.21256255

**Authors:** David M Presby, Emily R Capodilupo

## Abstract

Several vaccines against SARS-CoV-2 have been granted emergency use authorization from the United States Food and Drug Administration and similar regulatory bodies abroad to combat the COVID-19 pandemic. While these vaccines have been shown to be extremely safe, transient side-effects lasting 24-48 hours post-vaccination have been reported. Here we conducted a retrospective analysis of 50977 subscribers to the WHOOP platform (33119 males, 17858 females; total of 65686 unique responses) who received either the AstraZeneca (AZ, n=2093), Janssen/Johnson & Johnson (J&J&J, n=3888), Moderna (n=23776; M1, 14553 first dose; M2, 9223 second dose), or Pfizer/BioNTech (n=35929; P&B1, 22387 first dose; P&B2, 13542 second dose) vaccines using data collected through April 14, 2021. Subjective reactogenicity was assessed using self-reported surveys. Results from these surveys indicated that the odds of self-reporting an adverse event after vaccination depend on gender, age, and manufacturer. Objectively measured cardiovascular (resting heart rate, RHR; heart rate variability, HRV) and sleep (total sleep duration, % light sleep, and % restorative sleep [a combination of REM and slow wave sleep]) metrics were assessed using a wrist-worn biometric device (Whoop Inc, Boston, MA, USA) and compared to the same day of the week, one week prior. Data are presented as a percent change from baseline ± 95% confidence intervals. On the night after vaccination, RHR was higher (AZ: 13.5±0.76%; J&J&J: 16.5±0.64%; M1: 2.86±0.19%; M2: 9.3±0.53%; P&B1: 1.18±0.14%; P&B2: 13.5±0.36%) and HRV (AZ: -21.8±1.47%; J&J&J: - 25.6±1.15%; M1: -4.8±055%; M2: -19.9±1.33%; P&B1: -1.7±0.45%; P&B2: 8.60±1.10%) was lower than baseline levels. As for sleep metrics, total sleep was lower after the AZ and J&J&J vaccines (AZ: -3.7±0.98%; J&J&J: -3.8±0.80%; M1: 0.94±0.32%; M2: 0.14±0.80%; P&B1: 1.10±0.25%; P&B2: 0.35±0.63%); for AZ, J&J&J and the second dose of Moderna and P&B, a greater percentage of sleep post-vaccination came from light sleep (AZ: 9.24±1.22%; J&J&J: 13.8±1.02%; M1: 1.73±0.40%; M2: 8.02±0.99%; P&B1: 0.44±0.31%; P&B2: 2.54±0.74%) and a lower percentage from restorative sleep (AZ: -9.21±1.27%; J&J&J: -12.6±1.00%; M1: 0.16±0.43%; M2: -8.31±1.05%; P&B1: 1.27±0.34%; P&B2: -1.36±0.83%) than the week prior. Across all objective metrics measured, there were general trends that indicated an attenuated response in older populations and a larger response after the second dose for the Pfizer/BioNTech and Moderna vaccines (AstraZeneca second dose not analyzed). Importantly, the effects of the vaccines on cardiovascular and sleep measures were transient and returned to baseline by the second night following vaccination (P > 0.05 or absolute Cohen’s d < 0.25). In summary, these results confirm the previously observed subjective symptomatology trends, and for the first time show that objectively measured cardiovascular and sleep parameters are altered the night after vaccination. Moreover, these results suggest that the response may be different between vaccine manufacturers and may be modified by age and larger after the second dose. This information can be used to inform policy makers and employers considering offering paid time off for vaccination, as well as individuals planning their commitments post-vaccination.

## Introduction

Vaccines are needed to curb the spread of severe acute respiratory syndrome coronavirus 2 (SARS-CoV-2), which leads to coronavirus disease 2019 (COVID-19). Currently, vaccines from AstraZeneca, Janssen/Johnson & Johnson (J&J&J), Moderna, Pfizer/BioNTech have been approved for use across Europe and in North and South America^1^. While these vaccines effectively reduce SARS-CoV-2 infection^2–5^, initial reports on reactogenicity indicate that local and systemic events can occur transiently upon vaccination^6^. Previous data on reactogenicity relies on self-reported, subjective data and the effects of COVID-19 vaccine on objective physiological measures are unknown. As subjective data is prone to bias^7^, there is a need for objective data collection that can provide insights into additional dimensions, such as sleep architecture, which individuals are not consciously aware of.

Wearables that continuously collect high-quality physiological data provide an opportunity to examine the effect of COVID-19 vaccination on sleep and cardiovascular measures at larger scales than would typically be possible in traditional clinical settings. Here we utilize objective data collected using the WHOOP Strap (Whoop Inc Boston, MA, USA), a validated^8^, wrist-worn biometric device to determine the physiological effects of COVID-19 vaccination. This study is the first large-scale study to quantify the impact of available COVID-19 vaccinations using objective cardiovascular and sleep outcome data.

## Methods

### Data Collection

To gather subjective information on the effect of the COVID-19 vaccine, WHOOP members were prompted via daily surveys delivered through the WHOOP mobile application inquiring if they had received a COVID-19 vaccine. If they indicated that they had received a vaccine, they were further asked to provide the date of the vaccination, the manufacturer of their vaccine, and if they experienced any symptoms after vaccination. The symptoms questionnaire allowed for multiple selections, and queried for experience of a fever, chills, muscle aches, headaches, or fatigue. Users were also allowed to report no adverse events and this option did not allow for multiple selections. As part of the WHOOP data analytics platform, measures of sleep duration, architecture, and quality, as well as markers of cardiovascular health such as resting heart rate (RHR) and heart rate variability (HRV) are measured each night. For this analysis, metrics leading into and following vaccination were collated with the self-reported vaccine and symptomatology data. Baseline metrics were taken from the same day of the week on the week prior to the vaccination; the same day of week was used in order to control for confounders related to weekday/weekend differences in sleep behavior and outcomes which have been previously reported in WHOOP members^9^. Inclusion criteria consisted of being over the age of 18, having a WHOOP membership at the time of solicitation, and self-reporting having received one of the four vaccines included in this analysis. The survey reported in this manuscript is ongoing and includes data that were collected by April 14, 2021.

The utilized data are the property of WHOOP, Inc. and the authors are employees of WHOOP, Inc. and therefore are permitted to access the data used in this analysis per the company’s terms of service. The information security committee of WHOOP, Inc. concluded that this study does not require approval by an ethics committee due to the use of appropriately anonymized data accessed without the inclusion of personally identifiable information.

### Statistical analysis

All data were analyzed using the R programming language (version 4.0.4). For the self-reported surveys, data were analyzed using a logistic regression model that included manufacturer, discretized age, and gender. Age was discretized by binning into 18-29, 30-39, 40-54, and 55 & older age groups. Reference level for groups with more than two factors were either the Moderna second shot or the 18-29 year old group. To determine the effect of the vaccine on sleep and cardiovascular parameters, post-vaccine measures were compared to the prior same day of week within an individual using mixed model ANOVAs. These models included vaccination status (whether a person was pre- or post-vaccination, with post-vaccination defined here as the sleep immediately following vaccination), manufacturer, and age, and were subset into whether they came from the first or second/single dose. The single dose J&J&J vaccine was modeled as a second dose, reflecting that it is a final dose. When interactions were observed, models were further dissected to test for either simple two-way interactions or simple simple main effects. Pairwise comparisons between baseline and post-vaccine measures were made using t-tests and multiple comparisons were corrected for using the Bonferroni method. Effects sizes were determined using Cohen’s d. Values beyond three interquartile ranges from the upper or lower quartiles were deemed outliers and excluded from analysis. Significance was set as P < 0.05.

## Results

### Self-Reported Adverse Events Post-Vaccination

In total, we received 65867 responses (Table 1) to the voluntary COVID-19 vaccination survey. Using the “without an adverse event” response as a surrogate marker for not experiencing any symptoms, we found significant differences in the odds of reporting an adverse event for manufacturer, dose, age, and gender. Compared to the second dose of Moderna, those who received AstraZeneca (OR=1.19, 95% CI = 1.04-1.36; P < 0.05), Pfizer/BioNTech first dose (OR=7.82, 95% CI = 7.31-8.37; P < 0.001), Pfizer/BioNTech second dose (OR=2.94, 95% CI = 2.74-3.17; P < 0.001), Moderna first dose (OR=4.90, 95% CI=4.57-5.26; P<0.001), and J&J&J (OR=1.48, 95% CI=1.33-1.65; P<0.001) vaccines were more likely to report being without an adverse event (Figure 1). Men (OR=1.47, 95% CI=1.41-1.52; P <0.001) were more likely than women to report not experiencing an adverse event (Figure 1). Compared to the 18-29 year-old age group, those between the ages of 30-39 (OR= 1.21, 95% CI=1.16-1.27; P<0.001), 40-54 (OR=1.52, CI=1.45-1.60; P < 0.001), and 55 & older (OR=2.27, CI=2.12-2.44; P < 0.001) were more likely to report not experiencing an adverse event (Figure 1).

**Table 1.**
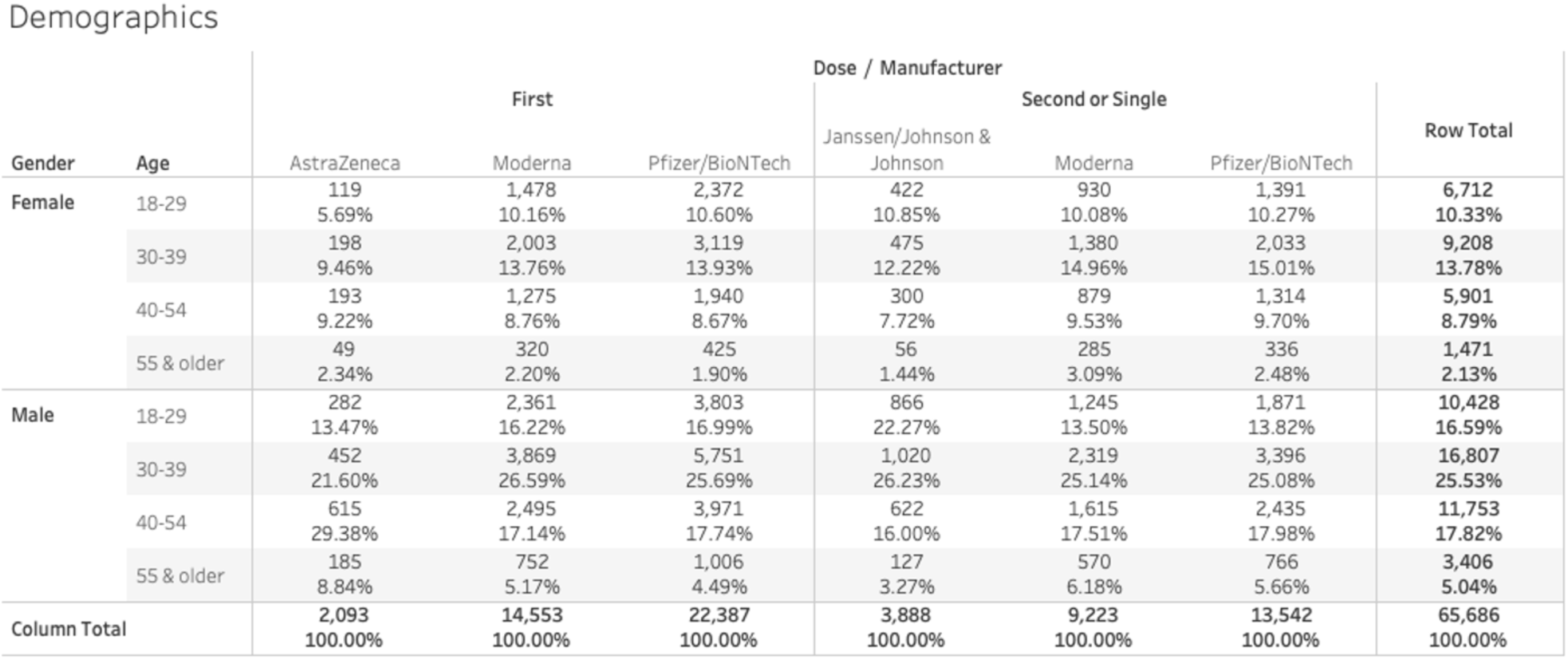
Participant demographics. Percentages reflect percent of column total.

**Figure 1.**
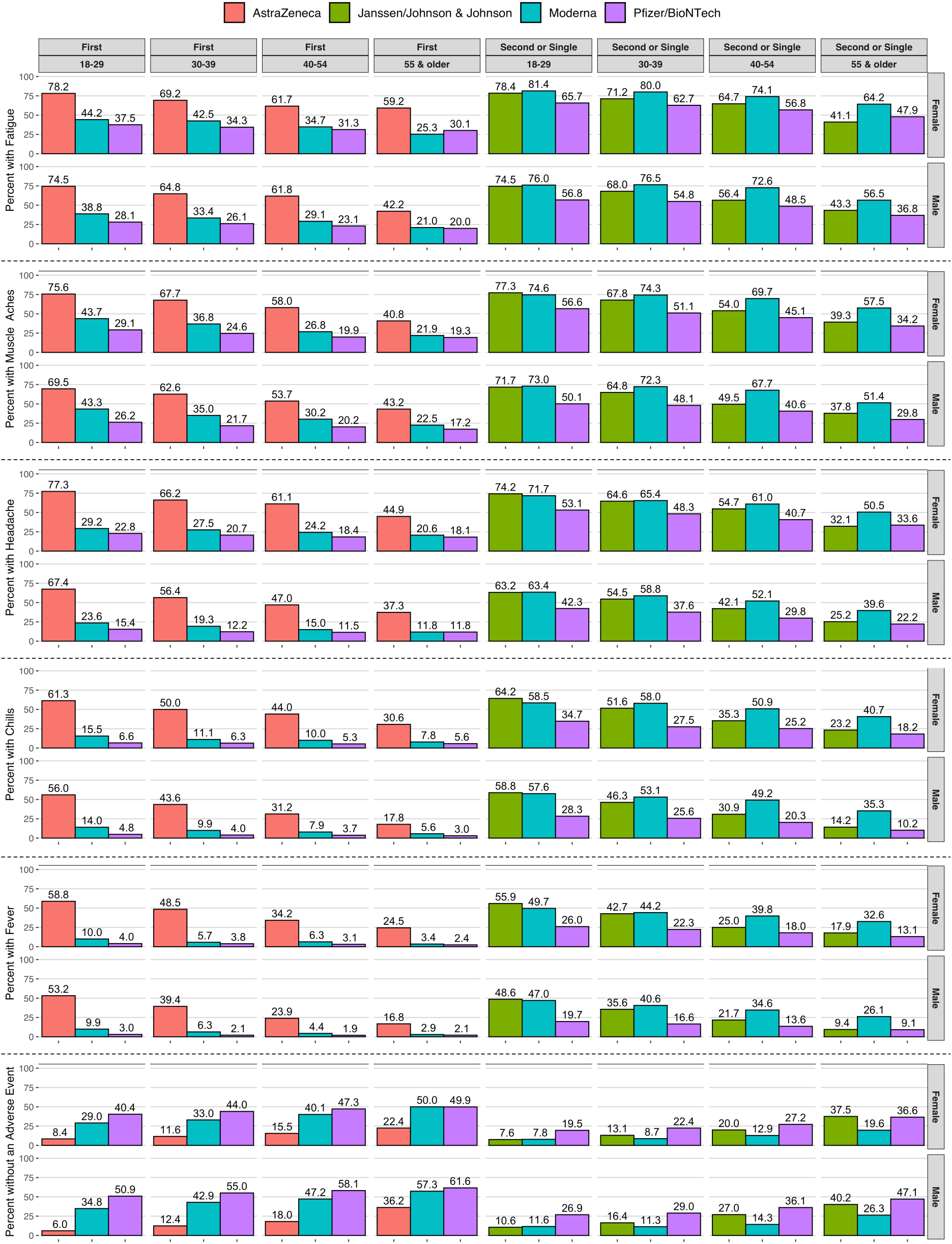
Self-reported Reactions After Vaccination. Participants were asked whether they experienced fatigue, muscle aches, a headache, chills, fever, or no side effects after vaccination. Data reflect percentage of responders within each cell for each vaccine (i.e. vaccine by sex, age group, and dose).

### Effect of COVID-19 Vaccination on Cardiovascular Parameters

Cardiovascular measures deviated from baseline values on the first night after vaccination (Figure 2A-D). Compared to individual baselines there was a three-way interaction between vaccination status, manufacturer, and age on RHR (P < 0.001; Supplemental Table 1) and HRV (P < 0.001; Supplemental Table 2) within each dosing regime. For both RHR and HRV, when subset by manufacturer, we observed interactions between vaccination status and age for each manufacturer (P < 0.001; Supplemental Table 1 and 2); when subset by age, we observed interactions between vaccination status and manufacturer for each age group (P < 0.001; Supplemental Table 1 and 2). We also analyzed the effect of the first vs second dose for the Moderna and Pfizer/BioNTech vaccines and found an interaction between vaccine status and first/second dose (P < 0.001; Supplemental Table 1 and 2). For RHR, pairwise comparisons and their associated effect sizes revealed that large effects were observed only in the younger individuals (<40 years old) and only after either the AstraZeneca, second Moderna, or J&J&J vaccination (Cohen’s d >0.8; Supplemental Table 3). Taken together, these results indicate that cardiovascular parameters are perturbed on the night after vaccination, and that these perturbations are modulated by age, manufacturer, and dose.

**Figure 2.**
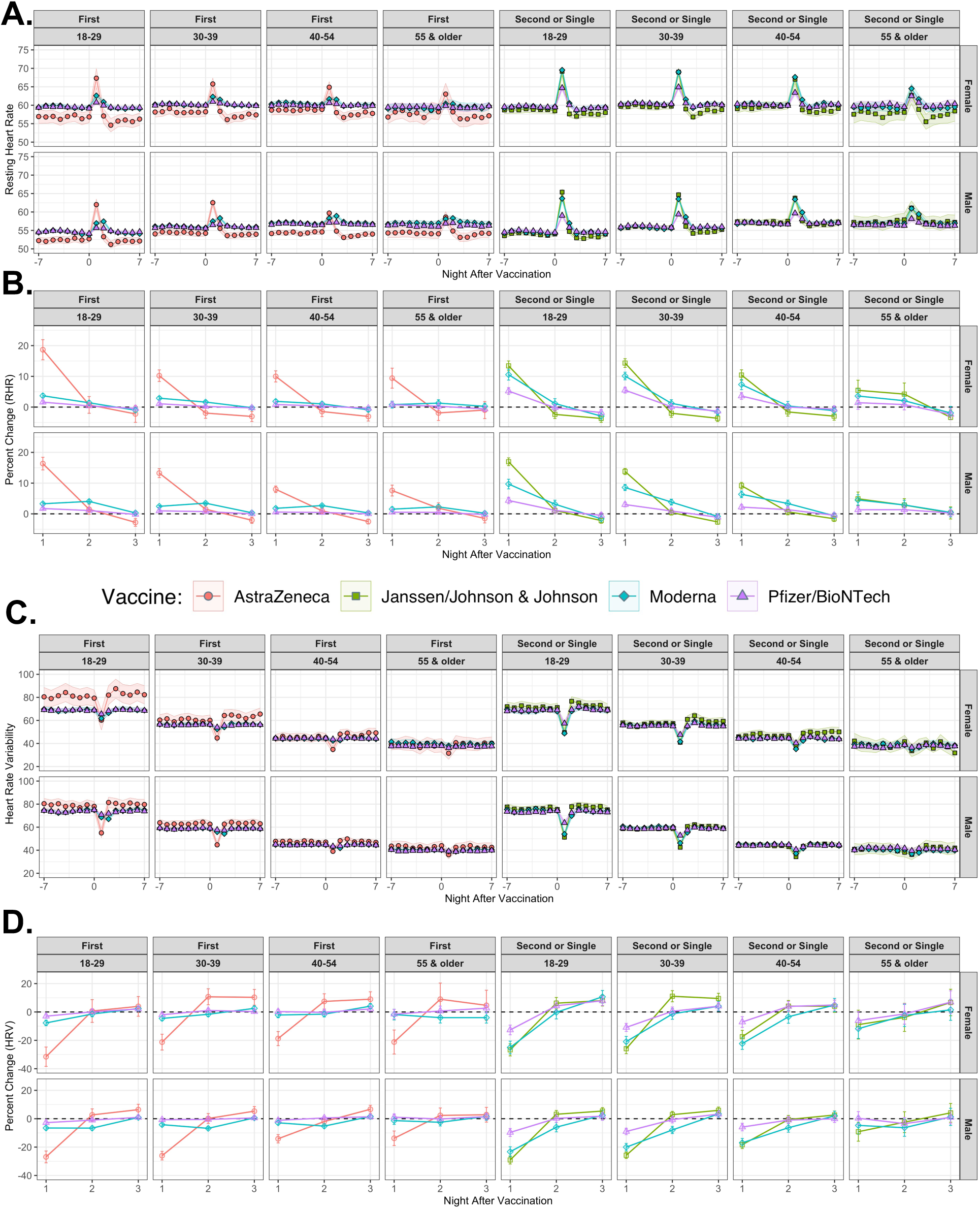
Cardiovascular Measures Before and After Vaccination. A) Resting heart rate (RHR) seven days before and seven days after vaccination. B) Percent change in RHR as compared to the same day of the week, one week prior. C) Heart rate variability (HRV) seven days before and seven days after vaccination. D) Percent change in HRV as compared to the same day of the week, one week prior. For all graphs, the title on the top indicates whether the data are from around the first, second, or single shot; the title on the second row indicates the age group. Data are presented as means ± 95% confidence intervals.

To determine the duration of the effect of the vaccines on cardiovascular parameters, pre-vs. post-vaccination pairwise comparisons were extended out to two- and three-nights post-vaccine; only Moderna and Pfizer/BioNTech vaccines were statistically significantly different from baseline on the second night in males. Specifically: on the second night after the first Moderna vaccine, HRV remained lower and RHR remained higher than baseline for members under the age of 55 (P < 0.001; Supplemental Table 4 and 5); on the second night after the second Moderna vaccine, HRV remained lower than baseline for members between the ages of 30-54 (all P < 0.001; Supplemental Table 5) and RHR remained higher than baseline for members under the age of 55 (all P < 0.001; Supplemental Table 4); on the second night after the second Pfizer/BioNTech vaccine, RHR remained higher than baseline for members between the ages 30-39 (P < 0.01; Supplemental Table 4). However, all of the differences observed on the second night were not meaningfully different from baseline (absolute Cohen’s d < 0.25; Supplemental Table 4 and 5). Together these results support the idea that cardiovascular parameters are affected by COVID-19 vaccination on the night after vaccination, and that these responses are both transient and modulated by age and manufacturer.

### Effect of COVID-19 Vaccination on Sleep Parameters

Total sleep and sleep architecture deviated from baseline values on the first night after vaccination (Figure 3A-F). For both the first and the second or single shot regimes, two way interactions were observed between vaccine status and age and between vaccine status and manufacturer on sleep duration (P < 0.001; Supplemental Table 6). Pairwise comparisons between baseline levels and the night after vaccination indicated that sleep is moderately decreased in the Male 18-29 year-old group that received the AstraZeneca vaccine (P<0.001, Cohen’s d<-0.5, Supplemental Table 7), while all other comparisons were either not significant or have negligible or small effects (absolute Cohen’s d <0.5; Supplemental Table 7).

**Figure 3.**
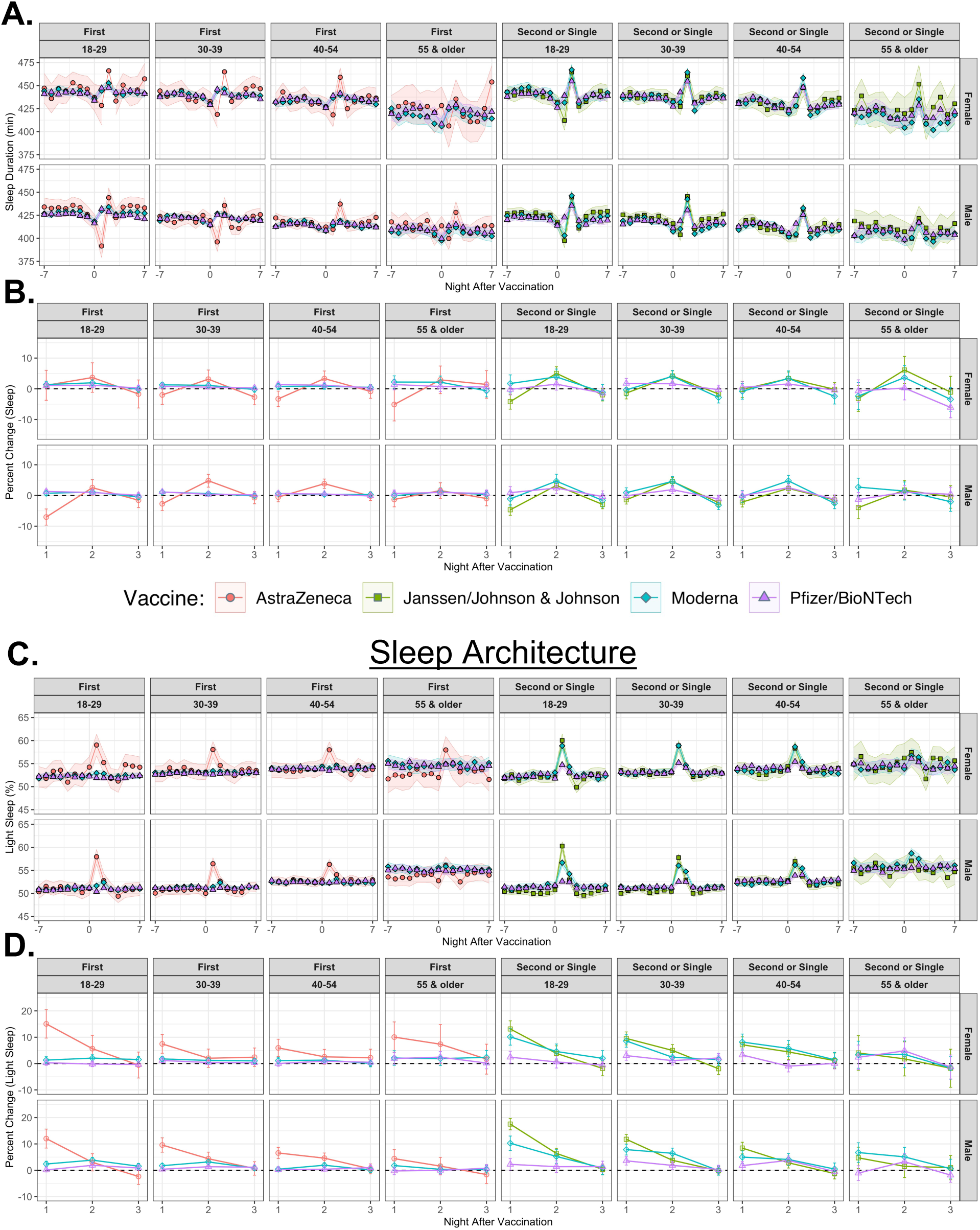
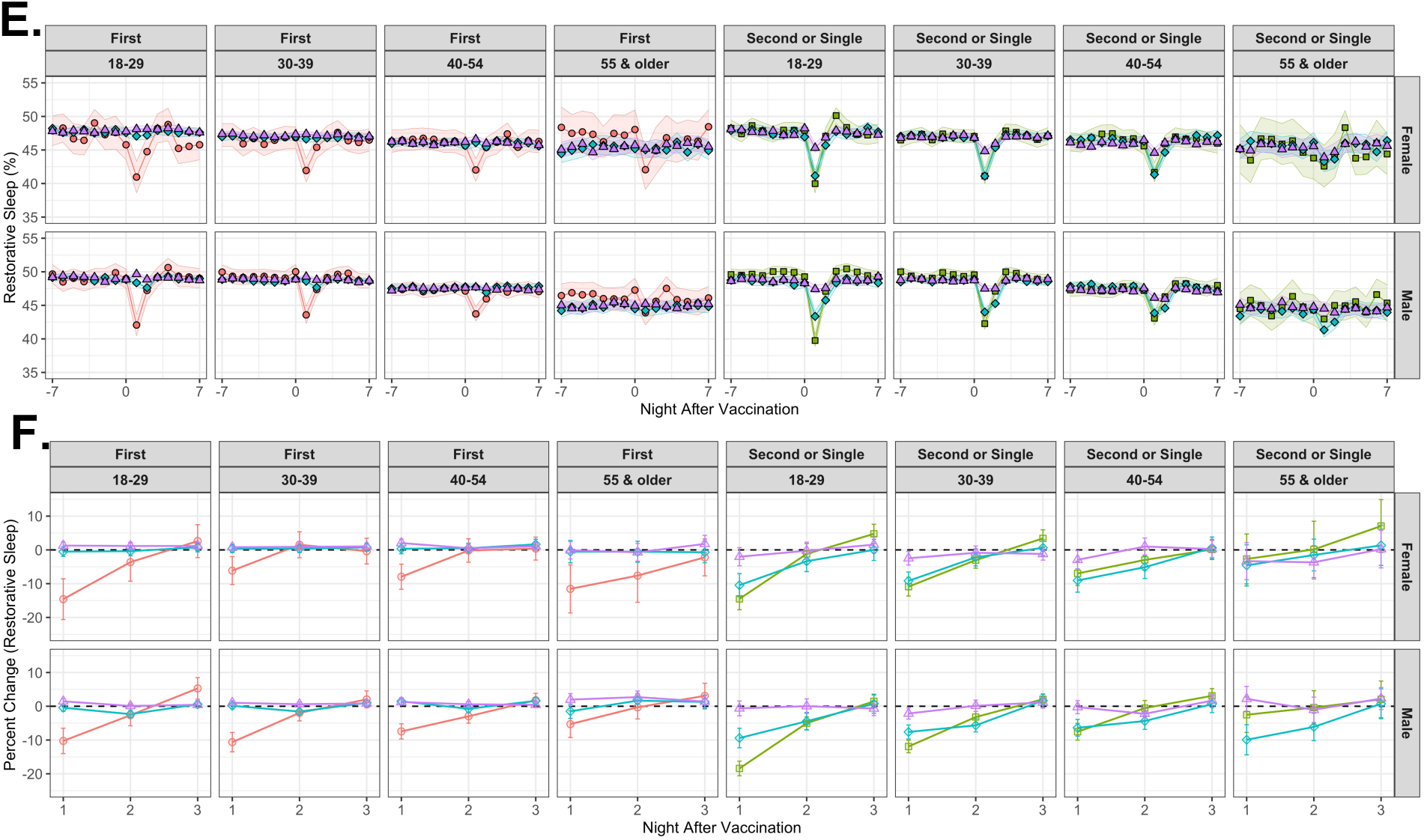
Sleep Metrics Before and After Vaccination. A) Total sleep duration (in minutes) seven days before and seven days after vaccination. B) Percent change in total sleep duration as compared to the same day of the week, one week prior. C) Percent light sleep seven days before and seven days after vaccination. D) Percent change in percent light sleep as compared to the same day of the week, one week prior. E) Percent restorative sleep seven days before and seven days after vaccination. F) Percent change in percent restorative light sleep as compared to the same day of the week, one week prior. For all graphs, the title on the top indicates whether the data are from around the first, second, or single shot; the title on the second row indicates the age group. Data are presented as means ± 95% confidence intervals.

For both light (P < 0.001; Supplemental Table 8) and restorative sleep (P < 0.001; Supplemental Table 9), there was a three-way interaction between vaccination status, manufacturer, and age within each dosing regimen. Pairwise comparisons and associated effect sizes indicated moderate effects on light or restorative sleep after the AstraZeneca, J&J&J, and second Moderna vaccine in individuals younger than 40 (P < 0.001, absolute Cohen’s d < 0.5; Supplemental Table 10 and 11), whereas the rest of the comparisons were either not significant or have negligible or small effects (absolute Cohen’s d <0.5; Supplemental Table 10 and 11). In summary, these results suggest that vaccines affect sleep and their associated components, and that these responses are modulated by manufacturer, dose, and age.

## Discussion

The aim of this study was to examine the objective and subjective response to COVID-19 vaccination in a large population of healthy adults. To accomplish this, we performed a retrospective analysis of the acute effect of vaccination on self-reported, sleep, and cardiovascular metrics on 50,977 individuals (providing 65,686 responses) receiving one of four COVID-19 vaccinations on or prior to April 14, 2021. Findings were divided into self-reported symptomatology, objectively measured sleep, and objectively measured cardiovascular metrics. The major finding of these analyses is that across the three sets of outcomes investigated, reactogenicity varied by vaccine dose (where applicable), manufacturer, and age of the recipient. These analyses complement and extend the initial findings from the individual clinical trials^2–5^ and self-reported surveys^6^ by comparing between more manufacturers and by examining objective measures. The results from this investigation suggest that the physiological response to COVID-19 vaccination, as assessed by cardiovascular and sleep measures, may differ by manufacturer and dose. This is the first study to investigate the effect of COVID-19 vaccination on cardiovascular and sleep parameters between manufacturers and dose and further research is required to determine the mechanisms behind these different responses.

In line with prior reports on subjective measures, we find that older vaccine recipients were less likely to report adverse events than younger participants^10^. We add to these reports by demonstrating that age also plays a role in the objectively measured physiological response to vaccination, whereby older individuals have an attenuated response as compared to younger participants across all objective metrics analyzed. It is well established that older individuals have less protection from disease after vaccination^11^ and we wonder if the attenuated responses measured within this investigation are due to an impaired immune response post-vaccination. If so, these results support a role for biometric monitoring to assess whether immunity was imprinted post-vaccine; however, further research needs to be conducted to address this hypothesis.

This is the first large-scale study to report on the acute objective and subjective reactogenicity of COVID-19 vaccination in healthy adults. These results indicate that cardiovascular and sleep parameters are disturbed on the first night after COVID-19 vaccination, and that the degree of disturbance is dependent on dose, manufacturer, and age. Importantly, the perturbations to cardiovascular and sleep parameters were transient and, in almost all analyzed cohorts, not practically different (Cohen’s d < 0.25) from pre-vaccine baseline levels by the second night following vaccination. These results are applicable to individuals anticipating receipt of a COVID-19 vaccine as they organize their post-vaccine commitments as well as by policy makers and employers considering offering time off for vaccination.

## Limitations

Interpretation of the results should be made with consideration to the limitations of this study. Data regarding the receipt of a COVID-19 vaccine, the preparation of vaccine received, and the symptoms experienced were collected via survey and not otherwise verified. However, we assume the frequency of recollection error was negligible due to the robustness of our findings and the short gap between vaccination and survey completion.

## Supporting information

RECORD CHECKLIST

## Data Availability

Public data sharing for these purposes is ethically and legally restricted. The data is collected and used with the consent of individuals who purchase a WHOOP membership (collectively, "WHOOP Members") and agree to the WHOOP terms of use (https://www.whoop.com/termsofuse/) and privacy policy (https://www.whoop.com/privacy/full-privacy-policy/). While WHOOP terms of use and privacy policy permit WHOOP to use collected data that has been aggregated or de-identified in a manner that no longer identifies an individual for informational purposes, analytics or WHOOP's own research purposes, the consent obtained from WHOOP Members does not extend to making the data publicly available for a third party to use for its own purposes. As such, WHOOP's legal department will not permit the data to be shared for these purposes.

## Contributions

DMP led the work of analyzing the data, DMP and ERC collectively designed the study, interpreted the results and wrote the manuscript.

